# Performance of readers and an artificial intelligence tool for grading of radiographic knee osteoarthritis at prespecified thresholds: Statistical analysis plan

**DOI:** 10.1101/2024.03.13.24304202

**Authors:** Mathias Willadsen Brejneboel, Mikael Boesen, Kay Geert A. Hermann, Edwin Oei, Huib Ruitenbeek, Katharina Ziegeler, Jacob J. Visser, Anders Lenskjold, Philip Hansen, Janus Uhd Nybing

## Abstract

1.

**Background and rationale:** Knee osteoarthritis (OA) is a common disease characterized by reduced function, stiffness, and pain. This clinical diagnosis is commonly supported with radiography of the weight-bearing knee. Radiographic features, such as the Kellgren-Lawrence (KL) grading system, are used as eligibility criteria for clinical studies while others, such as the OARSI grades and minimal joint space width, are used as endpoints for structural OA progression. A higher preoperative KL-grade has been correlated with better pain- and functional outcomes after knee arthroplasty surgery. Consequently, the KL-grade is a common requirement for approving knee arthroplasty among health insurance providers and it is commonly used by orthopedic surgeons as part of determining knee arthroplasty candidacy.

Historically, a radiologist was required to draw on and grade radiographs of the knee to extract the features. With increasing computational power and the increased use of deep convolutional neural networks, off-the-shelf artificial intelligence (AI) tools have become available for automatic extraction of these features. They have received regulatory approval for commercialization, but it is apparent that more diligent external validation is required. Finally, as AI tools begin to mature, new versions are released. It is important to assess how these developments change the current performance of the tool.

**Objectives:** The aim of this analysis is to evaluate the performance of a commercially available AI tool and of readers with different experience levels in orthopedic surgery and radiology at clinically relevant Kellgren-Lawrence grading system thresholds. Additionally, the performance of the AI tool for OARSI grades and patellar osteophytes will be evaluated across two versions of the AI tool.

**Methods:** This study is a secondary analysis of the data from the AutoRayValid-RBknee study, a retrospective observer performance study. It consists of non-fixed-flexion radiographs acquired from the production picture archiving and communications system (PACS) from three European centers. The primary outcome will be the difference in area under the receiver operating curve (AUC) between the readers and the AI tool at the prior authorization clinical criteria threshold (KL ≥ 3). Key secondary outcomes will be radiographic knee osteoarthritis (KL ≥ 2), osteoarthritis clinical trial inclusion (2 ≤ KL ≤ 3), and weight-loss trial inclusion (1 ≤ KL ≤ 3). The AUC of the readers will be computed using the SROC approach as proposed by Oakden-Rayner et al. Further, the performance of the AI tool for grading ordinal OARSI grades will be evaluated using the ordinal ROC as proposed by Obuchowski et al. and the AUC is used for estimating binary OARSI-grade and patellar osteophyte classification performance.

*Population:* Patients with knee pain referred for radiography on suspicion of knee osteoarthritis.

*Index test:* Readers
Each center will recruit four readers from across radiology and orthopedic surgery, one in-training and one board-certified for each specialty. AI tool
RBknee-2.2.0 (CE version, KL-grading, OARSI grading, patellar osteophytes) and RBknee-2.1.0 (CE version, KL-grading, OARSI grading, patellar osteophytes) will be used to perform the change impact analysis of advancing product development.

*Reference test:* The reference standard will be determined by the majority vote of three readers, one from each participating hospital who are a board-certified musculoskeletal radiology consultant with expertise in clinical and research evaluation of KOA including extensive experience using the KL-grade.

*Further statistical details:* Sample size
Not applicable as this is a secondary analysis. Framework
This is a diagnostic test accuracy study assessing the performance of a commercially available AI tool for radiographic evaluation of knee osteoarthritis according to established grading systems. Additionally, change impact analysis will be performed where multiple versions of the AI tool are available. Confidence intervals and P values
All 95% confidence intervals and P values will use an alpha of 5%. Multiplicity
No explicit multiplicity correction will be performed. Instead, a hierarchical approach will be taken based on tabular order of the tested hypotheses in Table 3. Statistical software
R version 4.2.2 (or newer).

## 2. ELABORATIONS ON OUTCOMES AND DATA

### Data management

Values outputted by the AI tool will be compared to the reference standard. Additionally, subgroup analyses will be based on image conformity.

#### Kellgren-Lawrence grade

On the frontal image (ordinal grades 0 = no osteoarthritis, 4 = severe osteoarthritis)[2].

#### Dichotomized Kellgren-Lawrence grade

The prior authorization clinical criteria threshold is set at KL ≥ 3, the radiographic knee osteoarthritis threshold is set at KL ≥ 2, osteoarthritis clinical trial inclusion threshold is set at 2 ≤ KL ≤ 3, and weight-loss trial inclusion threshold is set at 1 ≤ KL ≤ 3.[3,4]

#### OARSI grades

On the frontal image. Joint space narrowing (ordinal grades 0 = normal joint space, 3 = more than 2/3 narrowed) for the medial and lateral compartments, osteophytes (ordinal grades 0-3) for the medial and lateral femur and tibia and tibial eminence, subchondral sclerosis (yes/no) for the medial and lateral femur and tibia[5].

#### Patellar osteophytes

On the lateral image. Proximal and distal patellar osteophytes (yes/no).

#### Image nonconformity (not outputted by the index test)

Frontal images with rotation, angulation and/or inadequate for medial or lateral estimation (all binary). The groups are non-exclusive. An image can be both rotated and angled while only the lateral compartment is inadequate.

### Data validation

All variables used in the analyses, including the derived variables, will be checked for missing values, outliers, and inconsistencies.

### Data template

Based on this SAP, the statistical analyst will develop a tailored data template illustrating the data structure required for the statistical analyses.

## 3. OUTLINE

The analysis success rate of the AI tool will only be reported in the manuscript body.

The aggressiveness (absolute grading differences) between the current and previous versions of the AI tool will only be reported in the manuscript body.

The anticipated (predefined) outline of the manuscript is illustrated below.

### Further statistical information related to Table 1

Data will be presented as means with standard deviations (SD) when normally distributed or as medians with interquartile range in case of skewed data. Dichotomous and categorical data will be presented as absolute counts and proportions.

**Table 1.**
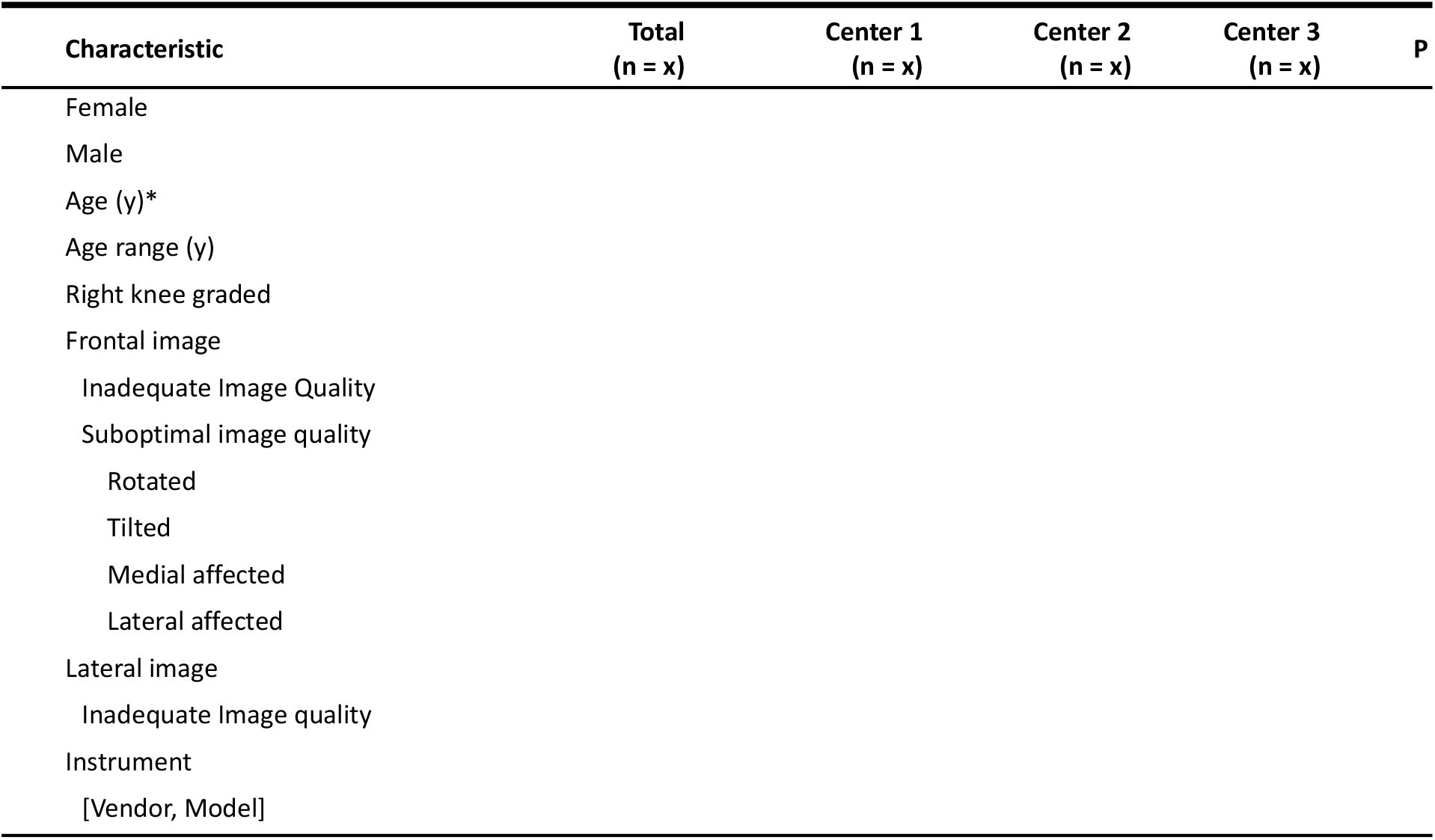
Patient characteristics. Values are reported as count (%) unless stated in the table. Inadequate image quality denotes images where the patient should be recalled for new imaging. Suboptimal image quality denotes images that would be used in the clinic, but which exhibit one or more suboptimal traits. JSW, Joint Space Width; OARSI, Osteoarthritis Research Society International; JSN, Joint Space Narrowing;

### Hypothetical ROC for the AI tool and SROC for the readers

The ROC of the AI tool and the SROC of the unassisted and AI assisted readers. The purple line is the ROC for the AI tool, the orange line is the SROC for the unassisted readers, and the blue line is the SROC of the AI assisted readers. The x-axis is 1 – specificity and the y-axis is sensitivity.

### Further statistical information related to Figure 2

The SROC of the readers will be used as proposed by Oakden-Rayner et al. treating each reader as a diagnostic test in a diagnostic test meta-analysis. The SROCs will be estimated using a mixed model approach using the Lehmann family as proposed by Holling et al. The diagonal dashed line from (0, 0) to (1, 1) indicates classification performance equal to chance.[6–9]

**Figure 1.**
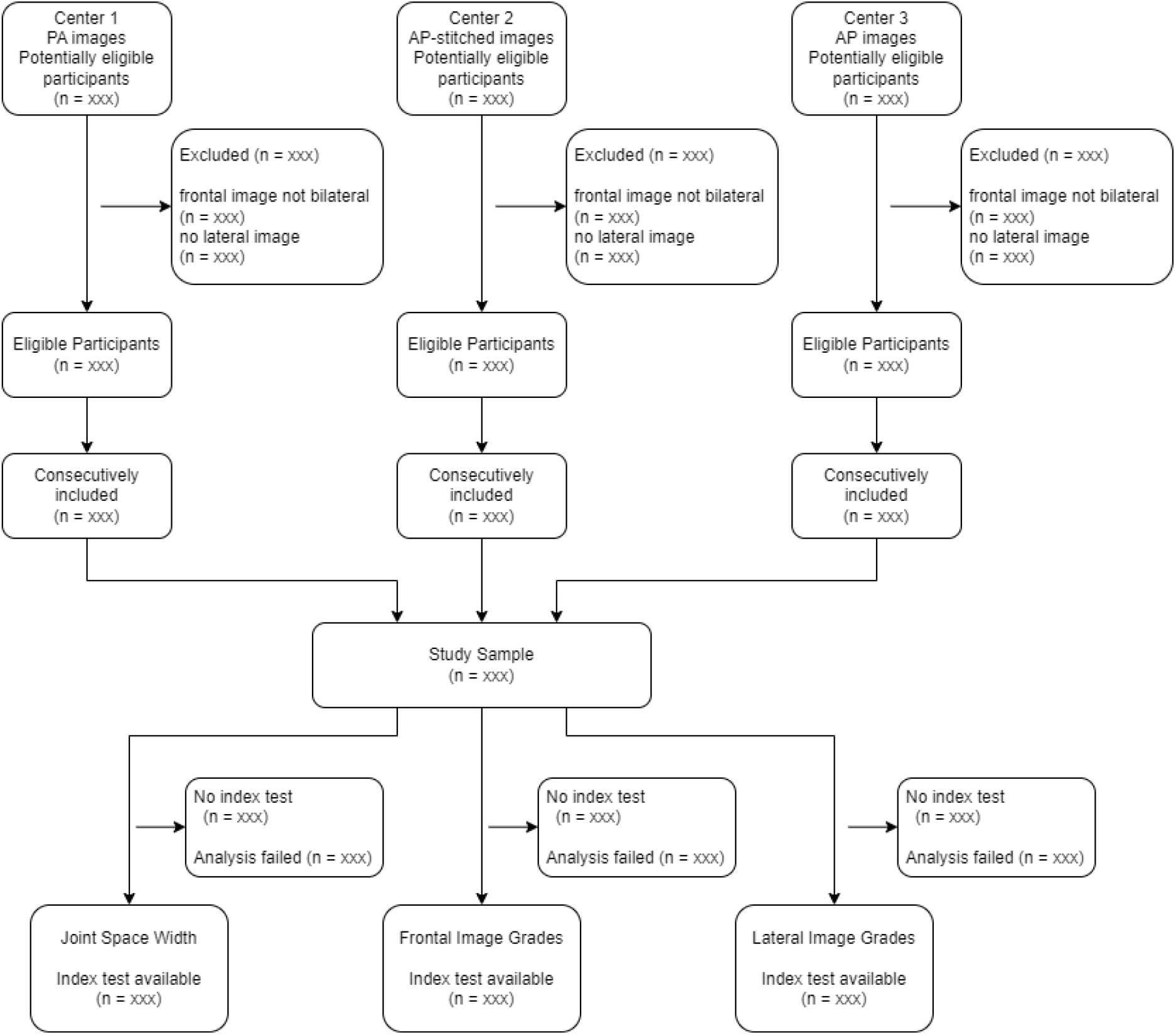
Flow diagram. *Anticipated plot design, illustrating potential reasons for exclusion:* PA: posteroanterior; AP: anteroposterior;

**Figure 2.**
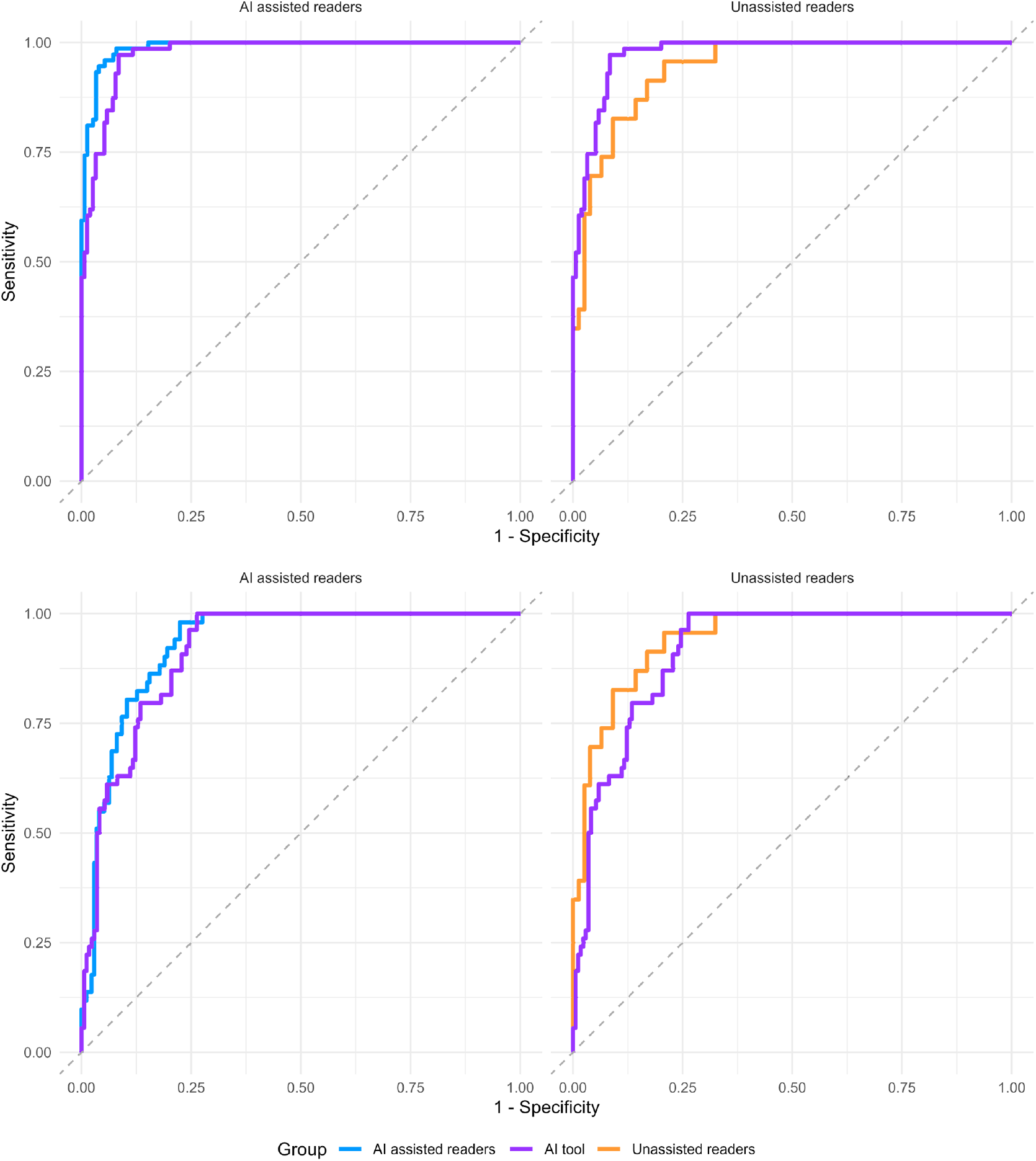
Receiver Operating Curve for classifying prior authorization clinical criteria (KL ≥ 3) and radiograpic knee osteoarthritis (KL ≥ 2) (MockUp)

### Further statistical information related to Table 2

Dichotomous and categorical data will be presented as absolute counts and proportions.

**Table 2.**
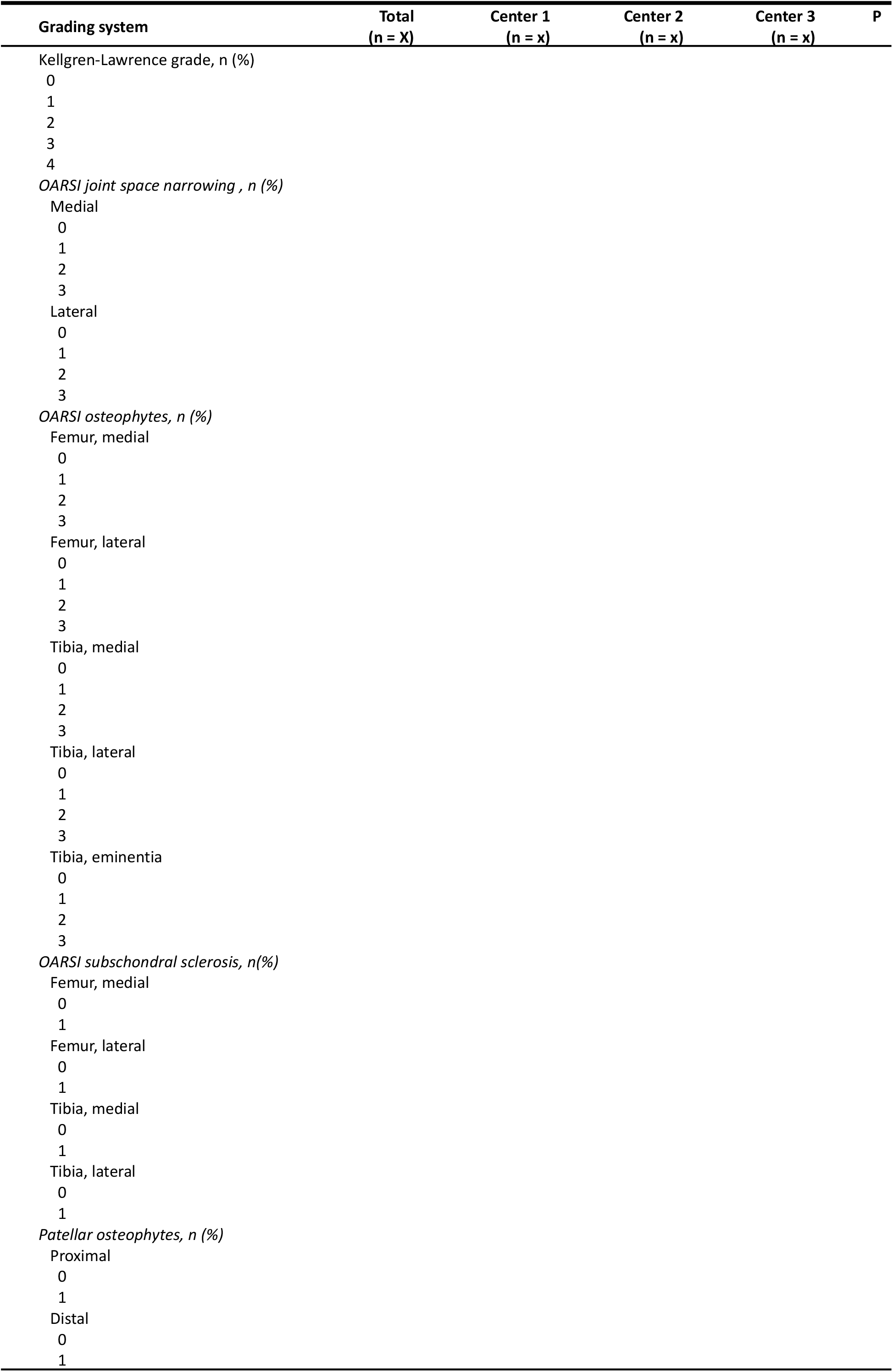
Grade distributions. Count (%) for frontal view Kellgren-Lawrence and OARSI grading systems, and lateral view patellar osteophytes. The *Chi*-squared test will be used to compare groups. OARSI, Osteoarthritis Research Society International.

### Further statistical information related to Table 3

The SROC of the readers will be used as proposed by Oakden-Rayner et al. treating each reader as a diagnostic test in a diagnostic test meta-analysis. The SROCs will be estimated using a mixed model approach using the Lehmann family as proposed by Holling et al. The ROC of the AI tool will be estimated as proposed by Robin et al. Groups will be compared using the *z* statistic from 10,000 bootstrap resamples as proposed by Robin at al.[6–9]

**Table 3.**
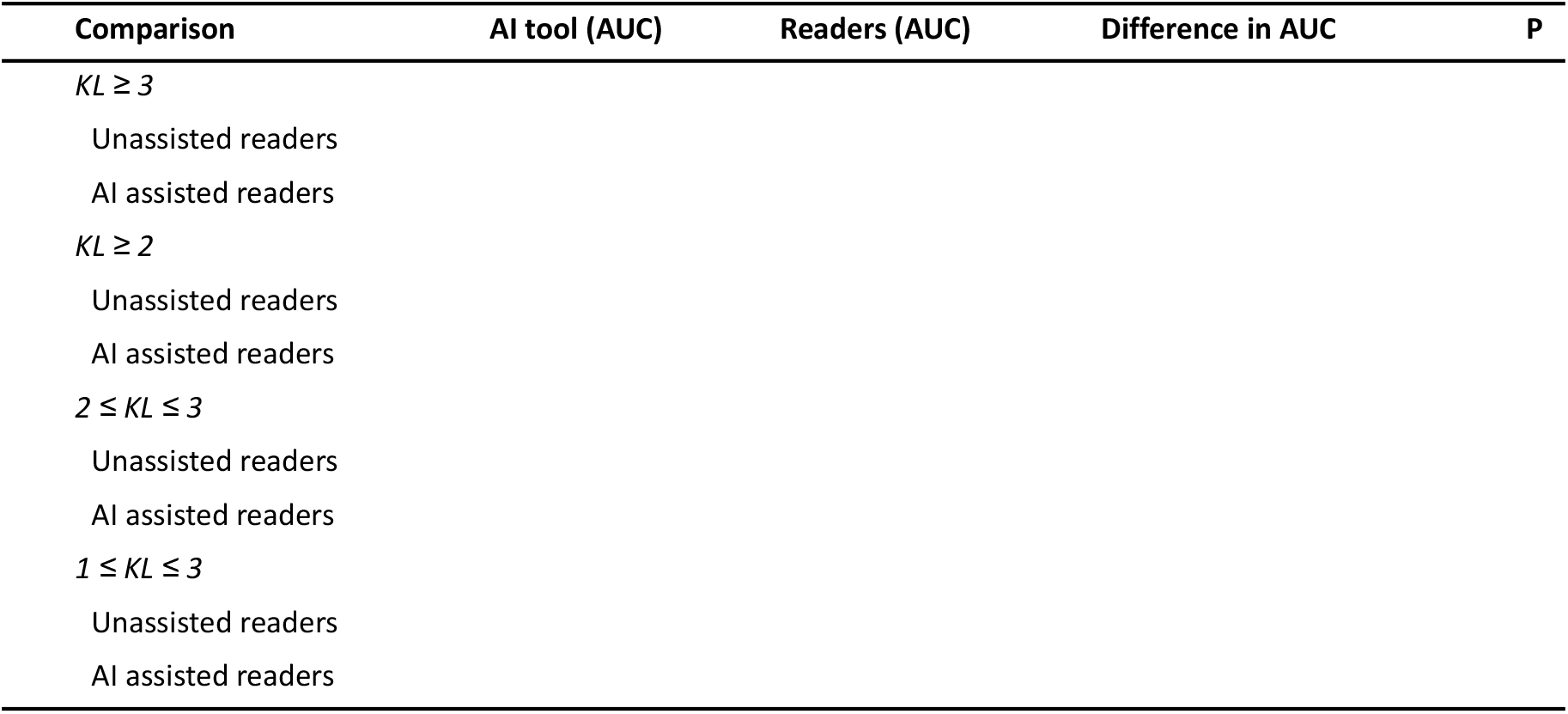
Comparison of the AI tool and the readers for dichotomized Kellgren-Lawrence grading at prespecified thresholds. Values will be reported as the AUC (95% confidence interval). Unassisted readers will refer to the readers without assistance from the AI tool and AI assisted readers to when the readers did receive AI assistance. OA, Osteoarthritis; OARSI, Osteoarthritis Research Society International; AUC, area under the receiver operating curve; SROC, summary ROC; ROC, receiver operating curve.

### Hypothetical ROC for the AI tool and SROC for the readers

The ROC of the AI tool and the SROC of the unassisted and AI assisted readers. The purple line is the ROC for the AI tool, the orange line is the SROC for the unassisted readers, and the blue line is the SROC of the AI assisted readers. The x-axis is 1 – specificity and the y-axis is sensitivity.

### Further statistical information related to Figure 3

The SROC of the readers will be used as proposed by Oakden-Rayner et al. treating each reader as a diagnostic test in a diagnostic test meta-analysis. The SROCs will be estimated using a mixed model approach using the Lehmann family as proposed by Holling et al. The diagonal dashed line from (0, 0) to (1, 1) indicates classification performance equal to chance.[6–9]

**Figure 3.**
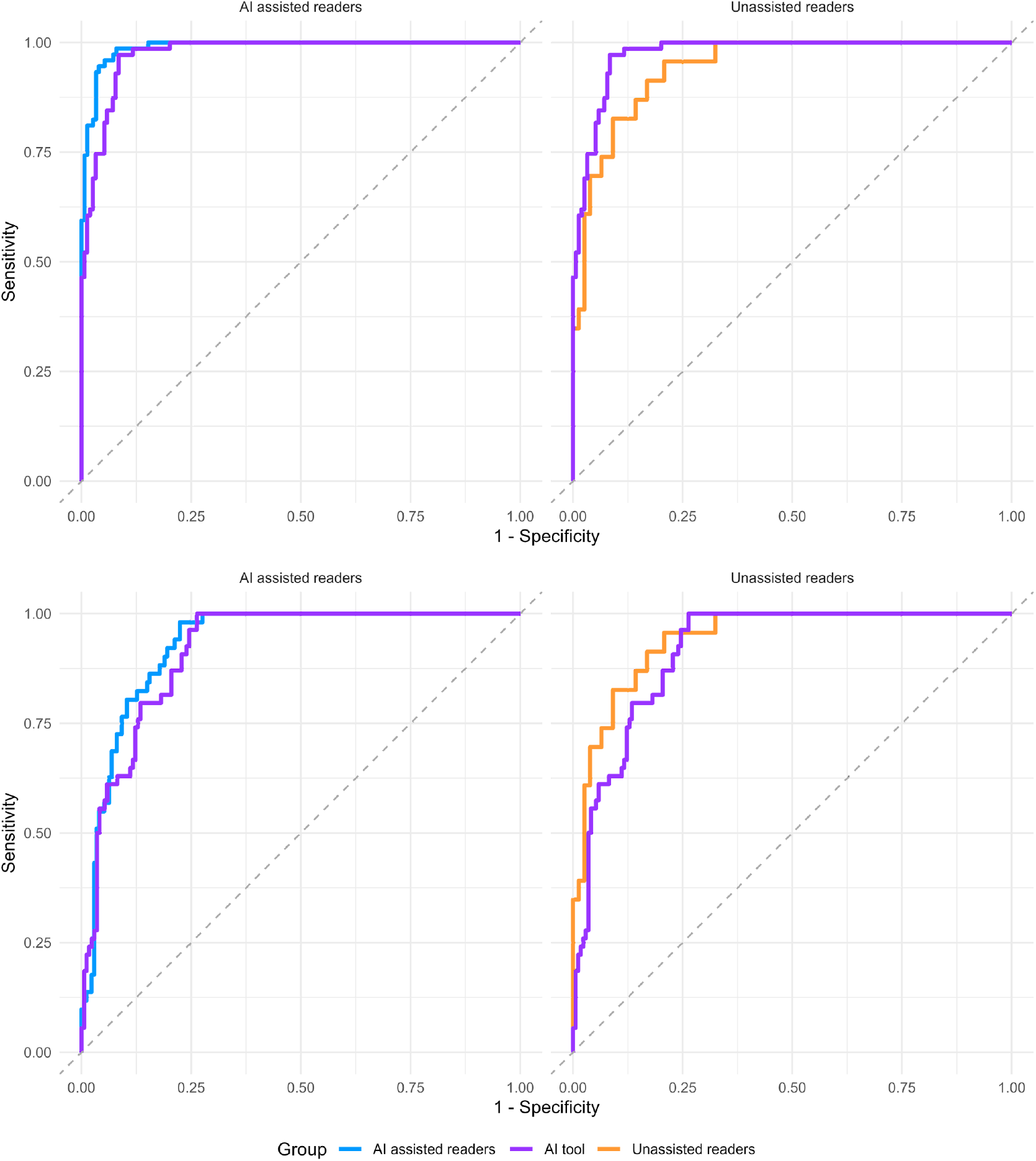
Receiver Operating Curve for classifying osteoarthritis trial inclusion (2 ≤ KL ≤ 3) and weight-loss related osteoarthritis trial inclusion (1 ≤ KL ≤ 3) (MockUp)

### Further statistical information related to Table 4

For ordinal variables, performance will be estimated as the AUC and compared across groups using the ordinal ROC method as proposed by Obuchowski et al.[10,11] For binary variables, performance will be estimated as the AUC of the ROC and groups will be compared using the DeLong method.

**Table 4.** Performance of the AI tool. Values will be reported as least squared means (standard error) unless noted otherwise in the table. Current refers to the newest version of the tested AI tool and previous refers to the second-to-newest version. OA, Osteoarthritis; OARSI, Osteoarthritis Research Society International; AUC, area under the receiver operating curve.

| Characteristics | Current | Previous | P |
| --- | --- | --- | --- |
| ***OARSI JSN (AUC):*** |  |  |  |
| Medial |  |  |  |
| Lateral |  |  |  |
| ***OARSI Osteophytes (AUC)*** |  |  |  |
| Femur Medial |  |  |  |
| Femur Lateral |  |  |  |
| Tibia Medial |  |  |  |
| Tibia Lateral |  |  |  |
| Tibia Eminence |  |  |  |
| ***OARSI Subchondral Sclerosis (AUC)*** |  |  |  |
| Femur Medial |  |  |  |
| Femur Lateral |  |  |  |
| Tibia Medial |  |  |  |
| Tibia Lateral |  |  |  |
| ***Patellar Osteophytes (AUC)*** |  |  |  |
| Proximal |  |  |  |
| Distal |  |  |  |

## Data Availability

All data produced in the present study are available upon reasonable request to the corresponding author.

## SUPPLEMENTARY MATERIAL

The anticipated (predefined) supplementary material of the manuscript is illustrated below.

**Supplementary file 1. Protocol**[1]

**Supplementary file 2. This SAP**

**Supplementary Table 3.**
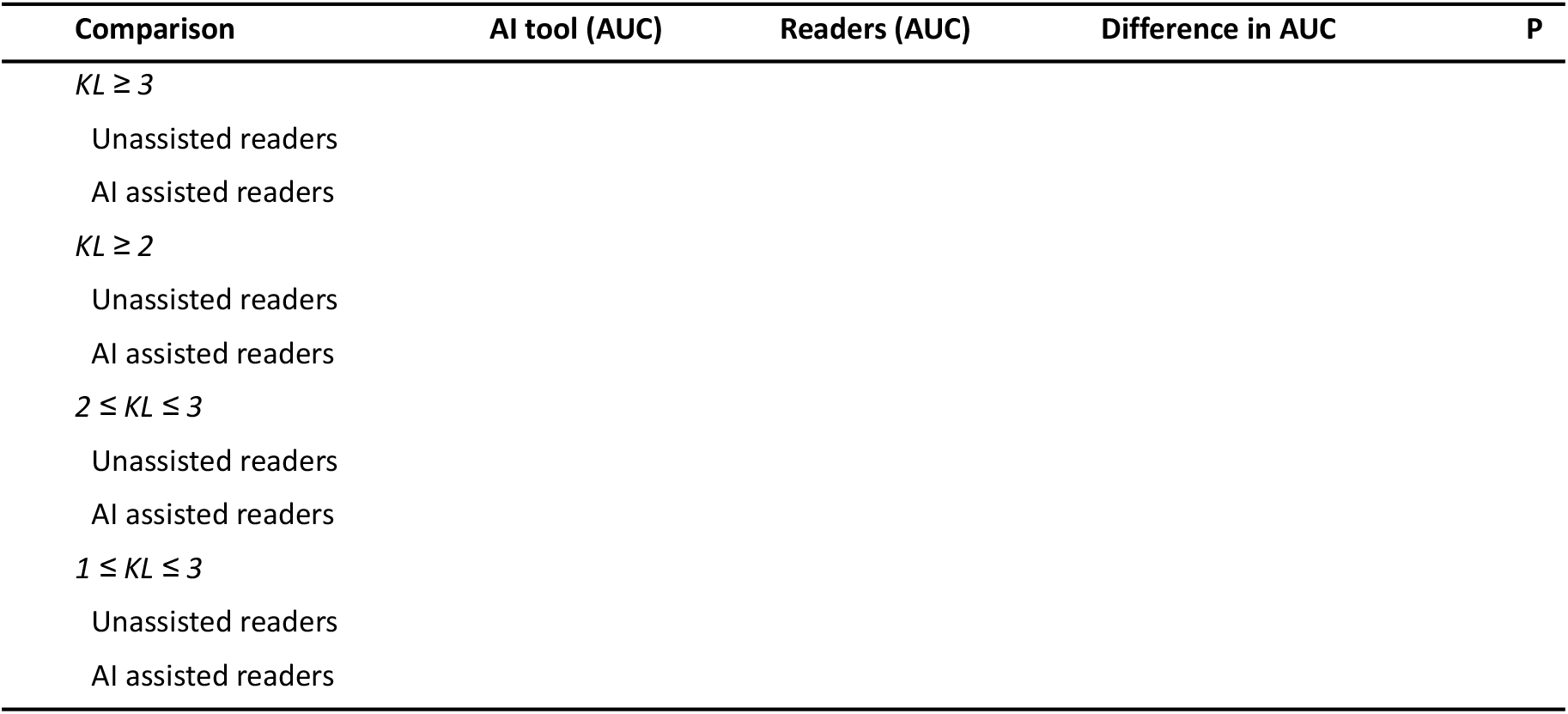
Comparison of the AI tool and the readers for dichotomized Kellgren-Lawrence grading at prespecified thresholds. Same as the primary analysis (Table 3) but only using images which were not considered of suboptimal quality. Values will be reported as the AUC (95% confidence interval). Unassisted readers will refer to the readers without assistance from the AI tool and AI assisted readers to when the readers did receive AI assistance. OA, Osteoarthritis; OARSI, Osteoarthritis Research Society International; AUC, area under the receiver operating curve; SROC, summary ROC; ROC, receiver operating curve.

### Further statistical information related to Supplementary Table 1

**Supplementary Table 2.**
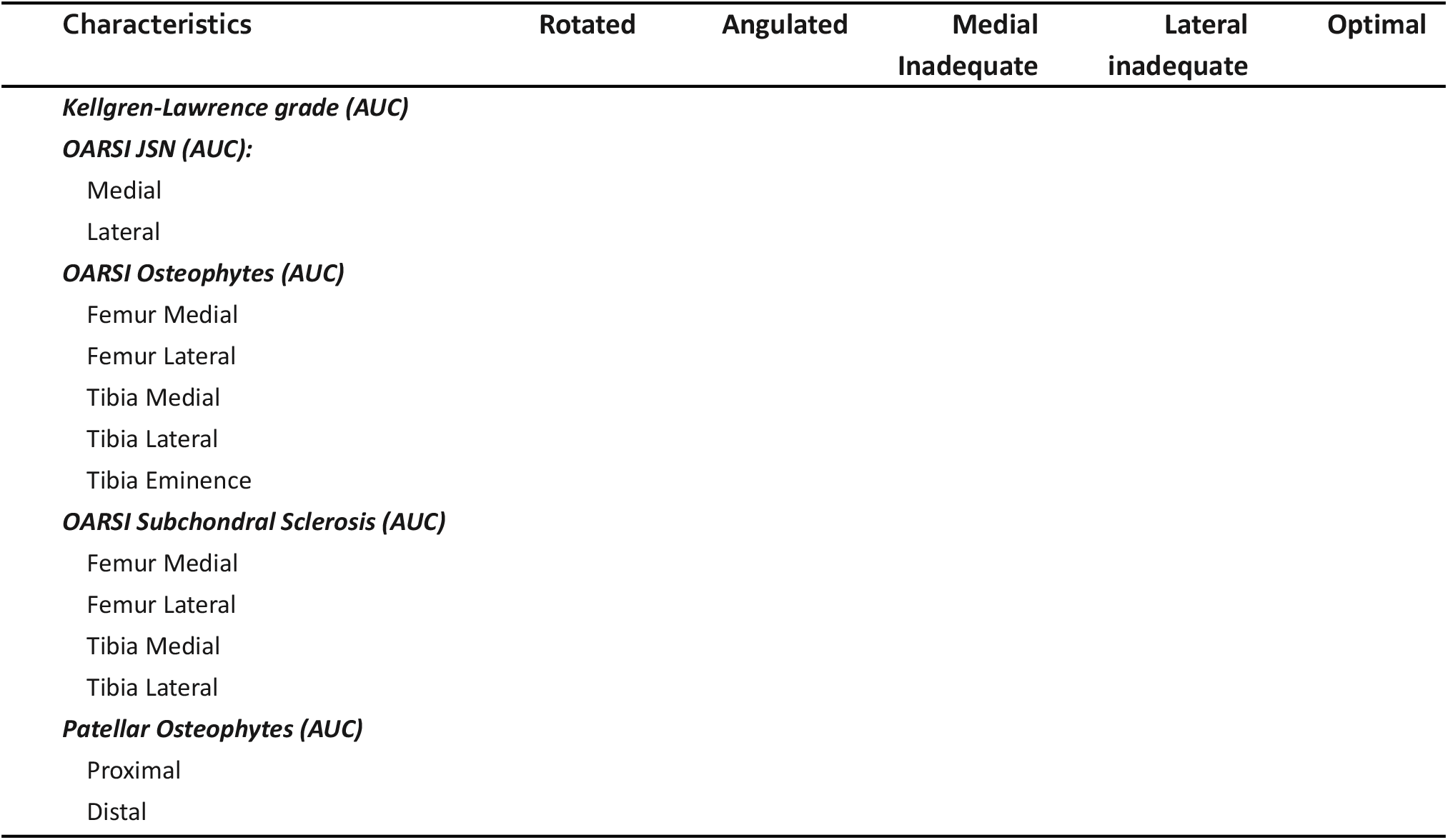
Performance of the AI tool for image nonconformity subgroups. Values will be reported as least squared means (standard error) unless noted otherwise in the table. OA, Osteoarthritis; OARSI, Osteoarthritis Research Society International; ord.acc, accuracy when the reference standard is ordinal; AUC, area under the receiver operating curve.

## 5. SAP REPORTING GUIDELINE

This SAP has been reported according to the items recommended in STARD guidelines by Cohen et al.[12] Explanation and elaboration of the items are available in the appendix paper.[13]

The guideline checklist and motivation is reproduced below.

| Section & Topic | No | Item | Reported on page # |
| --- | --- | --- | --- |
| **TITLE OR ABSTRACT** |  |  |  |
|  | **1** | Identification as a study of diagnostic accuracy using at least one measure of accuracy (such as sensitivity, specificity, predictive values, or AUC) |  |
| **ABSTRACT** |  |  |  |
|  | **2** | Structured summary of study design, methods, results, and conclusions (for specific guidance, see STARD for Abstracts) |  |
| **INTRODUCTION** |  |  |  |
|  | **3** | Scientific and clinical background, including the intended use and clinical role of the index test |  |
|  | **4** | Study objectives and hypotheses |  |
| **METHODS** |  |  |  |
| *Study design* | **5** | Whether data collection was planned before the index test and reference standard were performed (prospective study) or after (retrospective study) |  |
| *Participants* | **6** | Eligibility criteria |  |
|  | **7** | On what basis potentially eligible participants were identified (such as symptoms, results from previous tests, inclusion in registry) |  |
|  | **8** | Where and when potentially eligible participants were identified (setting, location and dates) |  |
|  | **9** | Whether participants formed a consecutive, random or convenience series |  |
| *Test methods* | **10a** | Index test, in sufficient detail to allow replication |  |
|  | **10b** | Reference standard, in sufficient detail to allow replication |  |
|  | **11** | Rationale for choosing the reference standard (if alternatives exist) |  |
|  | **12a** | Definition of and rationale for test positivity cut-offs or result categories of the index test, distinguishing pre-specified from exploratory |  |
|  | **12b** | Definition of and rationale for test positivity cut-offs or result categories of the reference standard, distinguishing pre-specified from exploratory |  |
|  | **13a** | Whether clinical information and reference standard results were available to the performers/readers of the index test |  |
|  | **13b** | Whether clinical information and index test results were available to the assessors of the reference standard |  |
| *Analysis* | **14** | Methods for estimating or comparing measures of diagnostic accuracy |  |
|  | **15** | How indeterminate index test or reference standard results were handled |  |
|  | **16** | How missing data on the index test and reference standard were handled |  |
|  | **17** | Any analyses of variability in diagnostic accuracy, distinguishing pre-specified from exploratory |  |
|  | **18** | Intended sample size and how it was determined |  |
| **RESULTS** |  |  |  |
| *Participants* | **19** | Flow of participants, using a diagram |  |
|  | **20** | Baseline demographic and clinical characteristics of participants |  |
|  | **21a** | Distribution of severity of disease in those with the target condition |  |
|  | **21b** | Distribution of alternative diagnoses in those without the target condition |  |
|  | **22** | Time interval and any clinical interventions between index test and reference standard |  |
| *Test results* | **23** | Cross tabulation of the index test results (or their distribution) by the results of the reference standard |  |
|  | **24** | Estimates of diagnostic accuracy and their precision (such as 95% confidence intervals) |  |
|  | **25** | Any adverse events from performing the index test or the reference standard |  |
| **DISCUSSION** |  |  |  |
|  | **26** | Study limitations, including sources of potential bias, statistical uncertainty, and generalisability |  |
|  | **27** | Implications for practice, including the intended use and clinical role of the index test |  |
| **OTHER INFORMATION** |  |  |  |
|  | **28** | Registration number and name of registry |  |
|  | **29** | Where the full study protocol can be accessed |  |
|  | **30** | Sources of funding and other support; role of funders |  |

## STARD 2015

### AIM

STARD stands for “Standards for Reporting Diagnostic accuracy studies”. This list of items was developed to contribute to the completeness and transparency of reporting of diagnostic accuracy studies. Authors can use the list to write informative study reports. Editors and peer-reviewers can use it to evaluate whether the information has been included in manuscripts submitted for publication.

### Explanation

A **diagnostic accuracy study** evaluates the ability of one or more medical tests to correctly classify study participants as having a **target condition**. This can be a disease, a disease stage, response or benefit from therapy, or an event or condition in the future. A medical test can be an imaging procedure, a laboratory test, elements from history and physical examination, a combination of these, or any other method for collecting information about the current health status of a patient.

The test whose accuracy is evaluated is called **index test**. A study can evaluate the accuracy of one or more index tests. Evaluating the ability of a medical test to correctly classify patients is typically done by comparing the distribution of the index test results with those of the **reference standard**. The reference standard is the best available method for establishing the presence or absence of the target condition. An accuracy study can rely on one or more reference standards.

If test results are categorized as either positive or negative, the cross tabulation of the index test results against those of the reference standard can be used to estimate the **sensitivity** of the index test (the proportion of participants *with* the target condition who have a positive index test), and its **specificity** (the proportion *without* the target condition who have a negative index test). From this cross tabulation (sometimes referred to as the contingency or “2x2” table), several other accuracy statistics can be estimated, such as the positive and negative **predictive values** of the test. Confidence intervals around estimates of accuracy can then be calculated to quantify the statistical **precision** of the measurements.

If the index test results can take more than two values, categorization of test results as positive or negative requires a **test positivity cut-off**. When multiple such cut-offs can be defined, authors can report a receiver operating characteristic (ROC) curve which graphically represents the combination of sensitivity and specificity for each possible test positivity cut-off. The **area under the ROC curve** informs in a single numerical value about the overall diagnostic accuracy of the index test.

The **intended use** of a medical test can be diagnosis, screening, staging, monitoring, surveillance, prediction or prognosis. The **clinical role** of a test explains its position relative to existing tests in the clinical pathway. A replacement test, for example, replaces an existing test. A triage test is used before an existing test; an add-on test is used after an existing test.

Besides diagnostic accuracy, several other outcomes and statistics may be relevant in the evaluation of medical tests. Medical tests can also be used to classify patients for purposes other than diagnosis, such as staging or prognosis. The STARD list was not explicitly developed for these other outcomes, statistics, and study types, although most STARD items would still apply.

### DEVELOPMENT

This STARD list was released in 2015. The 30 items were identified by an international expert group of methodologists, researchers, and editors. The guiding principle in the development of STARD was to select items that, when reported, would help readers to judge the potential for bias in the study, to appraise the applicability of the study findings and the validity of conclusions and recommendations. The list represents an update of the first version, which was published in 2003.

More information can be found on http://www.equator-network.org/reporting-guidelines/stard.

## Notes

### Competing Interest Statement

One author, Mikael Boesen, is a medical advisor for and shareholder of Radiobotics ApS.

### Funding Statement

This project has received funding from the European Union's Horizon 2020 research and innovation programme under grant agreement No 954221 for the EIC SME Instrument project AutoRay. The work only reflects the authors' view and the European Commission is not responsible for any use that may be made from the information it contains.

### Author Declarations

Danish Patient Safety Authority of Denmark waived ethical approval for this work. The IRB of Charite Universitatsmedizin - Berlin waived ethical approval for this work. The IRB of Erasmus Medical Center waived the ethical approval for this work.

### Summary of Updates

Revision, 2024 May 27. Expanded knowledge on the clinical relevance of the prior authorization clinical criteria at the Kellgren-Lawrence threshold of KL ≥ 3 has led to a thorough revision of this SAP. As originally planned, we will still evaluate the current and previous versions of the AI tool for the full set of semi-quantitative grades that it can analyze. Additions focus on the above-mentioned clinical threshold. To better estimate the real-world value of the AI tool, we have added a comparison of the AI tool and the readers in the original AutoRayValid-RBknee study. This comparison will be based on the SROC approach proposed by Oakden-Rayner et al. where each reader is treated as a diagnostic test study in a meta-analysis. The KL ≥ 3 threshold will be the primary outcome of the study, with key secondary outcomes being other clinically relevant thresholds: KL ≥ 2 (radiographic knee osteoarthritis threshold),2 ≤ KL ≤ 3 (inclusion criteria in osteoarthritis trials), and 1 ≤ KL ≤ 3 (inclusion criteria in osteoarthritis-related weight-loss trials). We removed the analysis of the joint space width measurements. These will be delegated to future research where fixed-flexion images will be included as well.

